# How Can New TB Vaccines Be Effectively Introduced in Indonesia? Insights from Diverse Stakeholders

**DOI:** 10.1101/2025.11.11.25340047

**Authors:** Nina Dwi Putri, Ahmad Fuady, Nugroho Soeharno, Aqila Sakina Zafira, Margareta Sirena Valeria, Pratama Wicaksana, Poppy Yuniar, Mardiati Nadjib, Azhiim Yudha, Katherine A Thomas, Rebecca A Clark, Sri Rezeki Hadinegoro, Richard G White

## Abstract

**Background:** New adult tuberculosis (TB) vaccines are in clinical trials and may be licensed as soon as 2028. However, vaccine rollout requires addressing multiple contextual factors beyond clinical trial findings. This study explored stakeholders’ perspectives on the introduction a new TB vaccine in Indonesia.

**Methods:** We used a mixed-methods approach combining a stakeholder consultation in Jakarta (13/03/2025) among 28 participants with diverse expertise and follow-up interviews with two of those participants. Participants completed a structured questionnaire (via Slido®) consisting of closed- and open-ended questions, adapted into Bahasa. Questions covered factors influencing vaccine introduction, target populations, delivery strategies, regulatory considerations, and lessons from other vaccine programs. Quantitative data were analysed descriptively, and qualitative responses underwent thematic analysis.

**Results:** All participants agreed on the importance of TB vaccine introduction. Key concerns were minimum efficacy of 50% and comparative effectiveness against other interventions, such as TB preventive treatment. Most agreed that vaccine introduction should not depend on local manufacturing capacity, administration route, or dosage. Critical enablers identified were adequate funding, strong political commitment, and demand generation through public acceptance. Priority target populations included people living with HIV/AIDS, individuals with diabetes, household contacts of TB patients, adolescents, and healthcare workers. Major challenges highlighted were vaccine hesitancy, halal issues, misinformation, and limited healthcare worker knowledge. Opinions diverged on feasibility of rollout without IGRA testing. Regulatory and budgeting processes were cited as additional barriers.

**Conclusions:** Introducing new TB vaccines in Indonesia will be complex and concerns extends beyond efficacy. Its success will depend on coordinated strategies to define target populations, design tailored delivery approaches, address vaccine hesitancy, and navigate regulatory and financing challenges.

## Introduction

Tuberculosis (TB) continues to be a critical global public health concern. In 2023, 8.2 million people globally were newly diagnosed with TB, and 1.25 million people died due to the disease[1]. Indonesia contributed 10% of the burden and became the country with the second highest TB burden globally, with an estimated 1.09 million people diagnosed with TB and approximately 125,000 TB-related deaths in 2023. As one of the countries with the highest TB incidence and a significant contributor to the global TB burden, Indonesia plays a pivotal role in global TB control efforts. Among many potential biomedical and social interventions to reduce TB burden is TB vaccine delivery, which may help block TB infection transmission in the population and prevent the illness and death[2].

The World Health Organization’s (WHO) new TB Vaccine investment case emphasizes the considerable economic and public health advantages of developing new TB vaccines to help meet global TB elimination goals as the vaccines could substantially lower TB incidence and death rates[3, 4]. The UN High-Level Meeting on TB also identified vaccines as one of the key strategies to eliminate the disease[5–7].

Given that the current BCG vaccine is most effective at preventing severe disease in children,[8] and its protection wanes over time, there is a critical need for effective vaccines for adolescents and adults—two populations that contribute substantially to transmission. The development of new vaccines targeting these groups has the potential to prevent both infection and onward transmission. that can prevent infection and transmission[9]. The development of such vaccines requires support from a strong global commitment in mobilising funding reaching up to USD5 billion a year by 2027[7]. Currently, there are 16 TB vaccines in the pipeline, and six of them–BCG traveller, GamTBVac, Immuvac, MTBVAC, VPM1002, and M72/AS01E, are in advanced Phase 3.[10] The M72/AS01E Phase IIb clinical trial conducted in Kenya, South Africa, and Zambia demonstrated significant protection against active TB disease among individuals with latent TB infection with an efficacy of 50% (90% confidence interval: 12–71%) over a follow-up period of about three years[11]. Following the promising results, the Phase III trial of this M72 vaccine is currently underway in which Indonesia is actively participating through five local trial centres.

However, despite the progress in vaccine development, the introduction of such new TB vaccine needs to consider multiple factors, such as the political situation, financial constraints, health system readiness, vaccine’s efficacy and safety,[12] as well as its comparison to other TB prevention methods. Perspectives from multiple stakeholders are therefore pivotal for a successful introduction and implementation[12, 13]. As the Phase III M72 trial vaccine results are expected in 2028, it is plausible that it may be licensed and available by 2030. Early and strategic preparation for its implementation is essential to ensure a smooth and timely launch. In 2024, a workshop was conducted involving key stakeholders to discuss initial potential strategies for introducing the TB vaccine in Indonesia[14]. The workshop recommended actions to improve TB vaccine availability, accessibility, and acceptability. Beyond developing health and economic impact models, it emphasized the need for a national stakeholder engagement plan spanning product development to introduction, to ensure coordinated and effective integration of TB vaccine activities across national, regional, and global levels. To ensure effective implementation, particularly given Indonesia’s status as a high-burden country and more updates and progress, strong evidence is needed to support the development of a model for TB vaccine rollout by involving a broader consultative meeting with more relevant stakeholders.

This study aimed to explore the perspectives of multiple stakeholders on the potential and challenges of introducing a new TB vaccine in Indonesia, taking into account various contextual factors.

## Methods

### Study Design

We conducted a multi-stakeholder consultation meeting in Jakarta, Indonesia, on 13^th^ of March 2025, utilizing a mixed-methods approach that integrated both quantitative and qualitative data collection and analysis. This approach was chosen to capture a comprehensive understanding of diverse, multi-stakeholder perspectives related to the potential of TB vaccine introduction and concerns around its uptake and implementation by the Indonesian government. The consultation was held as an in-person group meeting, facilitated by two trained moderators (NDP and AF). To further enrich the findings and address follow-up questions that emerged during the meeting, additional in-depth interviews were conducted with two selected meeting participants in April 2025.

### Participants

A total of 28 stakeholders were purposely selected for interview, based on their relevant capacity in tuberculosis, vaccine, and health policy (See **Supplement T1**). The selection aimed to ensure representation across all critical stakeholder groups that would be involved in TB vaccination decision-making and implementation.

We invited potential participants to the consultation meeting via email. The invitation included a structured explanation of the meeting’s objectives and planned activities. Confirmation from participants were obtained by phone, email, or direct messaging, and the actual enrolment when they provided informed consent was on 13/03/2025.

### Data Collection

At the start of the stakeholder meeting, participants received an overview of the TB disease burden, vaccine development, introduction pathways, and prior vaccine introduction experiences to ensure a shared understanding of the topic. Data were collected during the in-person one-day meeting through a structured individual questionnaire administered to all participants, consisting of both closed-ended (multiple-choice) items for quantitative analysis and open-ended questions designed to elicit detailed qualitative feedback, deployed in an online platform (Slido^®^). The questionnaire was firstly developed in English by the research team and Clinton Health Access Initiatives (CHAI) to be applied in several countries, e.g., India, South Africa, Brazil, Indonesia and it was translated and adapted to Bahasa (Indonesian Language) prior to the meeting. We grouped questions into several sections: factors affecting TB vaccine introduction, target population, strategy to deliver the vaccine in each population, regulatory consideration, and experience from other vaccine implementation.

NDP and AF guided participants to fill the questionnaire point-by-point. When participants had questions or problems in filling the questionnaire, including the interpretation of the questions, moderators helped explain and clarify the questions. Moderators also allowed participants to give individual reflection and group dialogue during the meeting to enrich the perspectives.

To address questions that emerged or remained unanswered during the meeting, additional in-depth interviews were conducted with two selected stakeholders who had also participated in the meeting. These interviews were led by the NDP and took place within two weeks after the meeting.

### Data Analysis

We employed a mixed-methods analytical framework. Quantitative data from the closed-ended questionnaire items were analysed descriptively and displayed as frequencies and proportions. Qualitative responses to the open-ended questions were subjected to thematic analysis. Two researchers (NDP and AF) independently reviewed and coded the responses, based on key themes/sections and subthemes/subsections in the questionnaire. Discrepancies in coding were discussed and resolved through consensus. Integration of quantitative and qualitative findings provided a nuanced understanding of stakeholder views. When relevant, we present participants’ quotes in the text and include their participant codes—explained in Supplement 1—in brackets at the end of each quote. For example: “response (*AAA1*).

## Results

Of 28 invitees, all attended the meeting. One participant left after the introductory presentation and did not complete the questionnaire at all. Although not all of the remaining 27 participants responded to every question, all available responses were included in the analysis. In total, there were a total 27 responses were recorded for this study. To address questions emerged or remained unanswered during the meeting, we interviewed two meeting participants through online interviews.

All participants agreed upon the importance of TB vaccine introduction given Indonesia’s high TB burden and the vaccine’s role in TB elimination although “it was not available yet” *(Participant code: GFM1)*. However, some concerns arose in the meeting.

### Factors affecting TB vaccine introduction

A summary of the most important factors affecting TB vaccine introduction is summarised in Figure 1. Of 26 participants responding to question on main factors affecting TB vaccine introduction decision, efficacy was perceived as the most important factor (n=22). Among those who cited efficacy as the main factor, 15 participants stated they would reject any vaccine with efficacy below 50%. When asked about the minimum efficacy required for introduction, all 21 respondents responding to this question agreed on at least 50% efficacy, with nine suggesting a higher threshold of 60–80%. Participants also perceived that the vaccine’s effectiveness related to other TB prevention strategies, including TB Preventive Therapy (TPT) (n=21), vaccination schedule (n=17), and the projected impact on the TB epidemic (n=17) would affect the decision for vaccine introduction.

**Figure 1:**
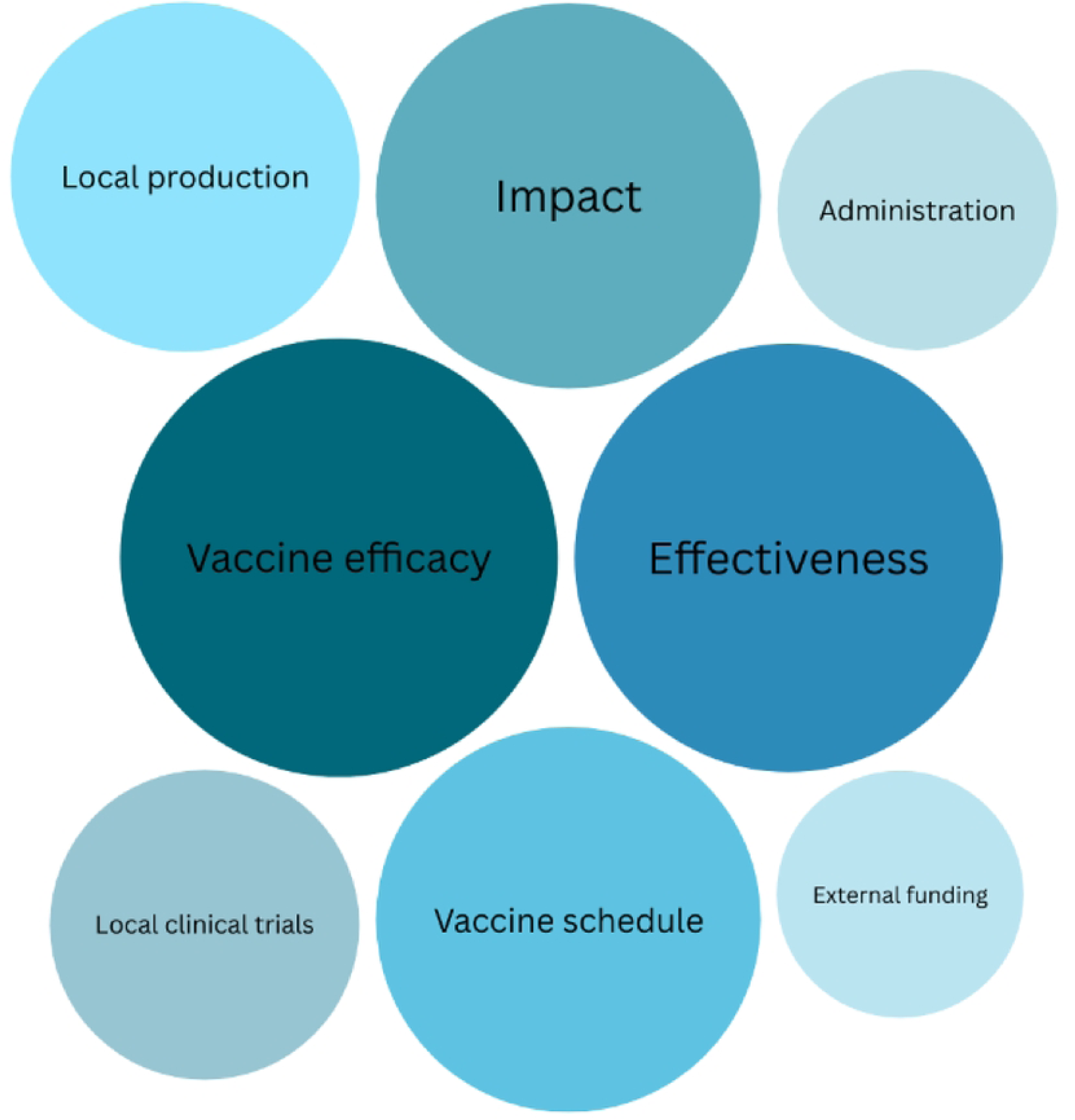
Factors influencing TB vaccine introduction.

Seventeen participants perceived that new TB vaccine would be more prioritized than TPT or other interventions, but some of them cited some conditions required, such as if vaccine efficacy >50% (n=7), proven long duration of protection (n=2), and less adverse events (n=1). There were different opinions among participants whether their preference of TB vaccine over TPT is applied in all populations or specific populations. However, despite this preference, a participant highlighted that TPT would be still needed “for those getting more benefits from TPT” (*XIM4*).

Most participants perceived that vaccine introduction would not depend on whether Indonesia has complete local manufacturing capability, the route of vaccine administration, and vaccine dosage. The dosage, instead, “would affect the coverage” *(GPL2)*, with single dose would be more “effective and efficient” *(XIM6)* “programmatically feasible” *(XIM5)* and “acceptable” *(XIM4)*.

Participants emphasized that, despite the benefit of TB vaccine, financial barriers could be constraints for TB vaccine introduction since “adequate funding is required for all aspects of TB vaccine uptake” *(XIM1)*. When asked about a new TB vaccine with 50% efficacy and 10 years of protection, participants expressed differing opinions on acceptable pricing. In general, lower prices were associated with higher acceptability, even two participants argued that “the vaccine should be free of charge” (*ETB7*). Price acceptability was influenced by multiple factors, including vaccine efficacy “too pricey for efficacy 50%” (*EPL1*), uncertainty about the 10-year protection, and national budget capacity, as reflected in the comments, “government is expected to cover [the price]” (*ETB9*) and “it would depend on national budget availability” (*GPL2*).

The issue of budget capacity could be overcome through strengthened political commitment, including strategic advocacy to governing political parties. Another challenge is the demand creation. Although the burden is high and prevention is required, the demand–the needs from people on TB vaccines–are required to be created by improving people’s understanding of the importance of prevention. It needs “demand creation and community involvement” *(XIM2)* through promotion and education but should also address the supply side to respond to the high demand. Community involvement was, therefore, cited as a critical issue prior to the vaccine introduction.

### Target population

Among 26 participants responding to target population question, in total, greater proportion of participants perceived that the vaccine needs to be prioritized for high-risk groups, people living with HIV/AIDS, those with diabetes, and household contacts of TB patients (**Table 1**). Other populations identified were adolescents 12-18 years old (n=19), healthcare workers (n=17). Sixteen participants selected the general population, defined as adults aged 18-64 years, as the priority group for TB vaccine introduction. When looking at the top-ranked priority groups, people living with HIV/AIDS (n=9) and adolescents (n=8) were most frequently identified as the highest priorities. The anticipated coverage of a new vaccine among each risk group and expected timeline is summarised in Table 2.

**Table 1.** Ranked priority populations for new TB vaccine introduction in Indonesia.

| Target<br>population | Ranking |  |  |  |  |  | Sum | Total (weighed<br>by rank)* |
| --- | --- | --- | --- | --- | --- | --- | --- | --- |
|  | 1st | 2nd | 3rd | 4th | 5th | 6th |  |  |
| People living with<br>HIV | 9 | 5 | 4 | 4 | 1 | 1 | 24 | 102 |
| Household<br>contacts of TB<br>patients | 3 | 6 | 8 | 2 | 2 | 0 | 21 | 84 |
| Adolescents (12–<br>17 years) | 8 | 2 | 4 | 1 | 2 | 2 | 19 | 83 |
| People living with<br>diabetes | 2 | 4 | 5 | 4 | 2 | 4 | 21 | 69 |
| Adults (18–64<br>years) | 1 | 6 | 2 | 4 | 3 | 0 | 16 | 68 |
| Healthcare<br>workers | 3 | 3 | 3 | 1 | 4 | 3 | 17 | 66 |

\* 1<sup>st</sup> ranking equals 6, 6<sup>th</sup> ranking equals 1

**Table 2:** Suggested implementation strategies for the priority populations.

| Target population | Delivery strategy | Coverage target | Years to achieve coverage | Feasibility |
| --- | --- | --- | --- | --- |
| **People living with HIV** | Roughly evenly split between routine first, campaign first and in parallel | RI: Most commonly 90%, ranging from 50-100%<br>Campaign: Most commonly 80% ranging from 10-100% | RI: Most commonly 1-2 years, ranging up to 10 years<br>Campaign: Most commonly 1-2 years, ranging up to 10 years | Viewed by most stakeholders as a feasible group to reach |
| **TB household contacts** | Majority suggested routine immunisation first | RI: Most commonly 80% ranging from 40-100%<br>Campaign: Most commonly 80% ranging from 30-100% | RI: Most commonly 2-6 years, ranging from 1-10+<br>Campaign: Most commonly 1-2 years ranging up to 10 | Viewed by most stakeholders as a feasible group to reach |
| **Adolescents** | Roughly even split between routine first, campaign immunisation first, or run in parallel | RI: Most commonly 80%, ranging from 30%-90%<br>Campaign: Most commonly 85%, ranging from 30%-100% | RI: Most commonly 2-6 years, ranging from 1-10+ years<br>Campaign: Most commonly 3-5 years, ranging from 1-10 years | Viewed by most stakeholders as a feasible group to reach |
| **People with diabetes** | Most stakeholders suggested either a campaign first or parallel approach | RI: Ranges from 30-90%<br>Campaign: Most commonly 50%, ranges from 30-90% | RI: Most commonly 2-6 years, ranges 1-10<br>Campaign: Most commonly 1-2 years, ranges from 1-10 years | Viewed by most stakeholders *not* as a feasible group to reach |
| **Adults** | Majority of stakeholder suggested having a campaign immunisation strategy first | RI: Most commonly 80% ranging from 30-100%<br>Campaign: Most commonly 80% ranging from 20-100% | RI: Most commonly 2-6 years ranging from 1-10<br>Campaign: Most commonly 1-5 years, ranged up to 10 | Viewed by most stakeholders as a feasible group to reach |
| **Healthcare workers** | Majority of stakeholders suggested routine immunisation first | RI: Ranges 30-100%<br>Campaign: Most commonly 90%, ranges between 30-100% | RI: Most commonly 1-2 years, ranges from 1-10 years<br>Campaign: Most commonly 1-2 years, ranges from 1-10 years | Viewed by most stakeholders as a feasible group to reach |

Individuals living with HIV/AIDS were perceived as having “a high risk of TB infection” *(GPL1, ETB1, ETB7, GIM1),* and they would “receive the biggest benefit” of this vaccine introduction *(ETB8)*. Targeting this population to be first vaccinated would “be easy to reach” *(EPL1)* since “they could be identified through HIV/AIDS” *(XIM4)*. Within this population, participants perceived that the coverage would be as high as 90% through the routine immunization program and would be achieved within 2-6 years.

People with close contact with individuals with TB were prioritized by participants, who argued that this group is particularly vulnerable due to their frequent exposure. Participants emphasized that vaccinating close contacts is important to “prevent transmission” and should be prioritized, especially in the context of limited financial resources.

The adolescent population was chosen as this group is perceived as living in “productive age” *(GTB1)*, and vaccinating this population would prevent productivity loss. Vaccinating this population would “prevent future transmission” *(XIM2)*, particularly “when they become young adults whose the high risk of infection” *(ETB1)* and “living in working age” *(GPL3)* and would “reduce TB burden in the future” *(ETB9)*. There were different perspectives among participants on how much the coverage and when the coverage would be reached. One participant, from a policy background, perceived that this vaccination would only reach 30% coverage within two years, while another participant perceived that it could reach 80% coverage but would need longer than 10 years.

People with diabetes were also identified as a priority group due to the high prevalence of TB–diabetes comorbidity. Stakeholders suggested either a campaign first or parallel approach between campaign and routine immunisations. For a campaign approach an expected coverage of 50% was suggested, ranging from 30-90%. Participants who prioritized adults as the target population reasoned that TB is most prevalent in this group and that they represent the productive age population. Stakeholders suggested a campaign vaccination approach would be most appropriate for adults, but for both campaign and routine immunisation stakeholders commonly suggested coverage could be around 80%. Although healthcare workers were recognized as being at increased risk of TB infection, participants generally perceived their priority as lower compared to other groups. Most stakeholders suggested they would be routinely vaccinated and would be able to achieve a high coverage at 90%.

We also explored experiences from previous vaccine and health program deliveries that could inform the rollout of the new TB vaccine. Participants identified several relevant challenges, including rejection from certain groups driven by hoaxes and misinformation. The issue of vaccine halal status was also perceived as particularly important to address, as concerns around it could potentially heighten vaccine hesitancy.

### Strategy to deliver a new TB vaccine

Strategy to deliver the vaccine would depend on the target population. Among adolescents, participants perceived that the vaccine can be delivered through routine immunization programs and campaigns. The delivery can be integrated in the extended “immunization month in school (*Bulan Imunisasi Anak Sekolah*, BIAS)” (*ETB7*) and school-based HPV vaccination (*XIM3*). The introduction of new TB vaccine can learn from HPV vaccine roll out through schools. While HPV vaccine is mostly currently delivered in elementary school, TB vaccine rollout to adolescents can be delivered to students in middle and high school, and this requires “substantial involvement with local Education Offices” (*GPL3*). Public campaigns are perceived as an alternative strategy as “campaigns are very efficient for this target group” (*XIM1*), and involving social media influencer (*ETB5*) and peer group campaign (*GPL6*) would be beneficial. For individuals with HIV/AIDS, it can be integrated with existing HIV/AIDS program. In addition, two participants with high familiarity with vaccine roll out mentioned that the new vaccine need to be introduced “in stepwise” approach *(EIM2, GIM1)* because “it would relate to the financing, and infrastructure readiness, such as cold chain and human resources” (*XIM5*). One expert mentioned different perspective on the strategy that the new TB vaccine “is ideally delivered by geographical strata instead of reaching specific population.” *(XIM6)*

### Potential challenges

Drawing from experiences during the COVID-19 vaccine rollout, participants identified some potential challenges for the new TB vaccine introduction, including vaccine hesitancy, concerns about halal status, and spread of hoaxes and misinformation. These factors were perceived a major contributor that could lead to “(public) rejection of the vaccine” *(EIM1, ETB7)*. Lack of vaccine knowledge among healthcare workers was also cited as potential challenges.

Interferon-gamma (IFN-γ) release assays (IGRA) status was cited as another challenge of the introduction. The phase 3 trials will generate efficacy among individuals with IGRA positive, not IGRA-negative. In the absence of local evidence on the efficacy of the new TB vaccine among IGRA-negative individuals, nine participants opposed rolling out the vaccine to this group, while six participants were uncertain. However, three participants argued that screening with IGRA was perceived as “not necessary” and doesn’t make sense since it “would be more expensive than the vaccine itself” *(ETB8)*.

While participants had different opinions on IGRA tests prior to vaccination, rolling out the vaccine to individuals without any IGRA status remained questionable since the vaccine use would be off-label if given to those with IGRA-negative. Off-label use, defined here as delivering the vaccine other than that for which it has been officially approved, was considered as not possible, referring to “there is no regulation allowing off-label use” *(XIM2, GPL3)*. The off-label use would be possible as an emergency, similar to the COVID-19 vaccine during the pandemic *(ETB8)*. However, despite its high incidence in Indonesia, TB is “not considered as an emergency situation” *(ETB9)*. Data on efficacy and safety among individuals with HIV/AIDS, IGRA negative, and unknown IGRA status, were cited as necessary prior to the roll out.

Other potential challenges cited in the meeting and interview were regulatory and budgeting issues. Registration of all medical products, including vaccines, will take time, for which “the maximum period of assessment is 52 weeks” (*GPL2*). Special treatment for acceleration would not be possible. TB vaccine “would not be treated as COVID vaccination since TB is not a pandemic,” (*GPL2*) but is subject to be accelerated depending on the data and document completion.

TB vaccine introduction also requires budget arrangement, planned one years before its implementation. The arrangement needs data on “efficacy, feasibility, acceptability, and price” *(GPL7)*. With the expected publication date on efficacy data in 2028, at the earliest, the normal process would only allow the TB vaccine introduction in 2030.

## Discussion

This study highlights insights from experts, policy makers, and relevant national and international organization partners on the potential introduction of a TB vaccine, and it emphasizes the importance of considering multiple interconnected aspects addressed by multiple stakeholders. The insights include the availability and strength of vaccine efficacy data, identification of appropriate target populations, comparative effectiveness with existing TB interventions, optimal delivery strategies, engagement of multiple stakeholders, regulatory and registration pathways, and the complexities of the budgeting, estimating numbers of vaccine needed, and financing processes. The findings underscore the multifaceted nature of TB vaccine introduction and the need for a comprehensive, coordinated approach to ensure its successful and sustainable implementation.

Efficacy emerged as a primary concern among stakeholders regarding the potential introduction of a new TB vaccine. The most promising candidate to date, M72/AS01E, demonstrated an efficacy of approximately 50% in preventing active TB disease among IGRA+ individuals in a Phase IIb trial.[11] In contrast, other vaccines targeted to adolescents such as the HPV vaccine have shown far higher efficacy—exceeding 90% in preventing infection with HPV types 16 and 18,[15] which sets a high benchmark for new vaccine introductions. Nonetheless, the 50% efficacy of M72/AS01E may still provide substantial population-level benefits, particularly when compared to BCG revaccination, which failed to protect against sustained *Mycobacterium tuberculosis* infection in QFT-negative, HIV-negative adolescents[16]. Clear and compelling efficacy data are therefore critical in shaping policy decisions and public confidence in TB vaccine rollout, including to convince public acceptance.

Despite its potential approximately 50% efficacy and its promising benefits in reducing TB incidence, participants perceived that TB vaccine should be viewed as a complementary strategy rather than a replacement for existing interventions. Tuberculosis preventive treatment (TPT) remains a critical component of TB control, particularly for populations who derive the greatest benefit from it, such as people living with HIV/AIDS[17, 18]. As noted by participants, the target populations for vaccination and TPT may differ, necessitating parallel and integrated implementation of both strategies[19, 20]. Although the TPT coverage is still low, 19.2% in 2025,[21] maintaining TPT alongside vaccine introduction ensures continued protection for high-risk groups and optimizes the overall impact of TB prevention efforts[22]. This highlights the need for coordinated policy planning that considers the strengths and limitations of each intervention within a broader public health framework.

Stakeholders highlighted that one of the major challenges in introducing a new TB vaccine is securing sufficient and sustained financing, as budget constraints may hinder implementation. Participants expressed concerns about vaccine pricing and advocated for it to be “as low as possible”, noting that the government would need to secure long-term funding. Price negotiation was considered critical to maximize population coverage and ensure broad public health impact. Given the government’s limited fiscal capacity, a phased or stepwise scale-up approach has been proposed to balance resource limitations with gradual program expansion, allowing for learning and adaptation along the way[23]. Timely data on supply, cost, target populations, infrastructure, and funding is essential for planning the sustainable implementation. In addition, successful vaccine rollout will require strong political commitment, which must be fostered through strategic advocacy efforts targeting policymakers and governing political parties[24]. Without such high-level support, the integration of a new TB vaccine into national immunization programs may be delayed or deprioritized.

Stakeholders also suggested that demand for a new TB vaccine needs to be created and it will require proactive efforts to combat misinformation, hoaxes, and vaccine-related misperceptions that have historically contributed to low vaccine uptake in various settings[25]. Community engagement, including community, ethnic, and religious leaders, is therefore essential—not only through culturally tailored education and health promotion—but also by building trust and addressing concerns that fuel vaccine hesitancy[25]. On the supply side, systems must be prepared to respond to increased demand, including ensuring cold chain capacity and service delivery integration. Stepwise implementation, from few provinces before scaling up into nationwide implementation is preferred. Financial planning should be also conducted in parallel, including advocacy for more affordable vaccine pricing and leveraging existing delivery platforms such as those used for school-based vaccine programs to optimize cost-efficiency[26–29].

Regarding regulatory approval, future clinical trials of TB vaccines should include diverse populations, particularly IGRA-negative individuals and people living with HIV, to ensure broader indications and equitable access. Requiring IGRA testing prior to vaccination was seen to pose significant logistical and financial challenges, especially in resource-limited settings, due to the high cost and complexity of implementation. These considerations highlight the importance of designing vaccines and delivery strategies that are operationally feasible and financially sustainable in real-world contexts.

This was the second, follow-up study exploring the perspectives of diverse stakeholders on the potential and challenges of introducing a new TB vaccine in Indonesia after the first meeting in November 2024. The participants were all relevant to this topic and provided insights into the various aspects that need to be considered prior to introduction. The meeting was open for live discussion among participants, allowing them to argue and revise their answers after the discussion which contributed to the richness of individual responses.

However, this study has some limitations. Despite the various background of participants, they may have limited knowledge on vaccine, particularly new TB vaccine, and could not fully understand some technical terms, such as efficacy. Additionally, the workshop was conducted in limited time, and probing to a deeper exploration was limited. These findings also did not include perspectives from the general population. As such, there is a need for additional research among target populations to understand their views on receiving a novel TB vaccine. Gaining this insight into the likely acceptability and coverage rates among key populations would help inform more effective delivery strategies. Furthermore, since the vaccine was not available at the time of the meeting, participants responded to hypothetical scenarios, leaving many uncertainties regarding the real-world vaccine efficacy, targeted population, and vaccine price. As Indonesia is participating in the M72 vaccine trial at the time of the stakeholder engagement, the responses provided by participants might have been influenced by the ongoing trial. As these responses were specific to Indonesia, these findings are not generalisable to other settings.

## Conclusions

The introduction of new TB vaccines in Indonesia has a multifaceted nature. For a successful implementation, the introduction requires a comprehensive, coordinated approach and consideration of several aspects, including the availability and strength of vaccine efficacy data, identification of appropriate target populations, comparative effectiveness with existing TB interventions, optimal delivery strategies, engagement of multiple stakeholders, regulatory and registration pathways, and the complexities of budgeting and financing processes.

## List of abbrevations

BCG: Bacillus Calmette–Guérin
HPV: Human Papillomavirus
IGRA: Interferon-gamma (IFN-γ) release assays
TB: Tuberculosis
TPT: Tuberculosis preventive treatment
WHO: World Health Organization.

## Declarations

### Ethics approval and consent to participate

Ethical approval for this study was obtained from the Faculty of Public Health, Universitas Indonesia (No. Ket-54/UN2.F10.D11/PPM.00.02/2025) and the Research Ethics Committee of the London School of Hygiene & Tropical Medicine (LSHTM) (31510 /RR/37086). All participants received a written explanation of the study at the beginning of the meeting before providing their written informed consent to participate.

### Consent for publication

Not applicable.

## Availability of data and materials

Data are available upon request to the corresponding author. The request should be specific and will be assessed on a case-by-case basis by all authors. There are no personalised data in this study, but all data sharing will still abide by rules and policies defined by the involved parties. Data sharing mechanisms will ensure that the rights and privacy of individuals participating in research will be always protected.

## Author Contributions

Conceptualization, NDP, AF, NS, RGW; Methodology, NDP, AF, NS, ASZ, RC, RGW; Software, ASZ, MSV, PW; Validation, NDP, AF, NS, PY; Formal Analysis, NDP, AF, NS; Resources, NS, PY, MN; Data Curation, NDP, AF, NS, ASZ, MSV, PW, PY, MN, AY; Writing – Original Draft Preparation, NDP, AF; Writing – Review & Editing, NDP, AF, NS, KAT, RGW; Visualization, NDP, AF, KAT; Supervision, SRH, RGW; Project Administration, ASZ, MSV, PW, PP, AY; Funding Acquisition, NS, SRH, RGW.

## Acknowledgments

We acknowledge the support of Inke Maris and the Clinton Health Access Initiative for their assistance in organizing the meeting.

